# Persistent transmission of Schistosomiasis in Northwest Nigeria: A community-based assessment of urogenital and female genital infections

**DOI:** 10.1101/2025.08.01.25332782

**Authors:** S. I. Yelwa, M. M. Dogara, J. B. Balogun, Y. A. Kani, S. S. Dawaki, A. K. Adeniyi, A.A. Ibrahim, A. U. Abdurrahaman, D. Ahmed

## Abstract

Urogenital schistosomiasis (UGS) and female genital schistosomiasis (FGS) remain significant public health concerns in Northwestern Nigeria, particularly among vulnerable populations like adolescent girls and women of reproductive age. Despite ongoing control efforts, transmission persists. This study investigated the prevalence, risk factors, and socio-demographic characteristics associated with UGS and FGS in selected areas of Jigawa State.

This study enrolled 648 females from six local government areas. Urine samples were analyzed for *Schistosoma haematobium* eggs to determine the prevalence of urogenital schistosomiasis (UGS). Additionally, 606 participants underwent gynecological examinations to assess the prevalence of female genital schistosomiasis (FGS) based on characteristic lesions. Bivariate and multivariate statistical models were used to analyze the relationships between socio-demographic data, water contact behavior, sanitation practices, and the prevalence of UGS and FGS.

The study found an overall prevalence of urogenital schistosomiasis (UGS) as 13.6% (95% CI: 11.0–16.5%), with Auyo having the highest rate (6.3%) and Buji the lowest (0.6%). Heavy infections (≥50 eggs/ml urine) were most common in Ringim (57.1%) and Taura (50%). For female genital schistosomiasis (FGS), the prevalence was 25.1% (95% CI: 21.7–28.7%), with Buji (44.5%) and Dutse (38.7%) reporting the highest rates. Multivariate analysis revealed age and geographic location as significant predictors of infection (p < 0.001). Adolescents aged 10–14 years were at highest risk for UGS (OR = 5.56, p = 0.006), while older women were more likely to have FGS due to cumulative exposure.

This study highlights the transmission dynamics of UGS and FGS in relation to socio-demographic factors in Jigawa State’s irrigation-dependent areas. To combat the disease, we recommend ongoing interventions, including mass drug administration (MDA) with praziquantel, improved sanitation, health education, and enhanced diagnostics. Targeted control measures should prioritize high-risk groups, especially adolescents and women, to reduce disease burden and break transmission cycles.

## INTRODUCTION

Schistosomiasis is an acute and chronic disease caused by dioecious blood flukes (trematode) of the genus *Schistosoma*. There are two major types of disease: intestinal schistosomiasis caused by *S. mansoni*, *S. japonicum*, *S. mekongi*, *S. guineensis*, and *S. intercalatum* and urogenital schistosomiasis, caused only by *S. haematobium*. All species have a specific snail as their intermediate host (1). Each of these species has a tropism for different body organs, presenting with genitourinary, intestinal or systemic disease, while the two minor species: *S. mekongi* and *S. intercalatum* are tropic for the intestines and liver (2). The disease remains one of the most neglected tropical diseases (NTDs) with severe public health implications, particularly in endemic regions of sub-Saharan Africa, including Northwest Nigeria (3).

The disease, caused by parasitic worm of the genus *Schistosoma*, primarily manifests as urogenital schistosomiasis (UGS) and female genital schistosomiasis (FGS); conditions that are frequently underdiagnosed and overlooked in endemic communities (4). UGS is commonly associated with *Schistosoma haematobium*, affecting the urinary tract and causing hematuria, fibrosis, and in severe cases, bladder cancer (5). On one hand, FGS, a less recognized but equally debilitating condition, is a result of *S. haematobium* eggs lodging in vaginal, cervical, and uterine tissues, leading to chronic inflammation, infertility, and increased susceptibility to HIV and other sexually transmitted infections (6). Therefore, infection with *S. haematobium* is notably associated with morbidity of the male and female genitals, bladder, and kidneys (7). This is because S. *haematobium* worms live in the veins that supply the bladder, uterus, and cervix, among other major pelvic organs.

Global Burden of Disease Study (8) and Hale Collaborators (9) reported that schistosomiasis was estimated to cause the loss of 1,440 million disability-adjusted life years (DALYs). Many women that acquired *S. haematobium* infection in childhood, about 30% to 75% of them may develop female genital schistosomiasis (FGS) (10, 11). The global prevalence of FGS is not known, but it has been reported to be high in poor and rural communities in the tropical and subtropical parts of the world, especially those which do not have access to adequate sanitation and safe water (11, 12, 13). However, Berne, (14) reported that infections with *S. haematobium* in women in endemic areas range from 33-75%. It is estimated that up to 56 million women in sub-Saharan Africa have FGS, and almost 20 million more cases will occur in the next decade unless girls are treated (13). Many believed that the burden is greatly underestimated (15, 16). The heavy burden was estimated 224 million suffer the malignant effects of the disease with an estimated 280,000 death toll every year mostly among the rural inhabitants (17, 18) while 90% population of in endemic communities are at risk of infections with schistosome infections (19, 20).

Regardless of age or gender, any demographic group that encounters polluted water in an endemic location where there is lack of access to clean water, poverty, ignorance, and poor hygiene habits is at risk of developing the disease. Additionally, communities engaged in anthropogenic activities such as swimming, farming, fishing, washing clothes and bathing in ponds, rivers, and dams where the intermediate snail hosts breed are at high risk of infection (21, 22, 23). Nigeria is the country with the highest prevalence of human urogenital schistosomiasis (24). It is perhaps the most important disease associated with man-made lake, irrigation projects in tropical countries (24, 25, 26).

The disease is reported from all the six geo-political zones in Nigeria (27), with about 29 million cases in 2008 (24, 28) and an estimated population of 101 million people are at risk (29). It is perhaps the most important disease associated with man-made lake, irrigation projects in tropical countries (24, 25, 26). There is evidence for a link between the occurrence of human urogenital schistosomiasis and several factors such as knowledge, attitude, perception, behavioral, cultural and religious practices, primary occupation, educational level, household income. All these factors go a long way to influence the transmission of urogenital schistosomiasis in any endemic settlement (17, 24, 25, 30). Northwest Nigeria remains one of the most endemic regions for urogenital schistosomiasis, particularly among school-aged children and women of reproductive age (31). Socio-environmental factors such as poor sanitation, limited access to clean water, and persistent human-water contact in infested bodies of water exacerbate the disease burden in Nigeria (32). Additionally, gender disparities in health-seeking behavior and cultural norms contribute to the underreporting of FGS cases, which is often misdiagnosed as other reproductive health conditions (33). Moreover, Jigawa State, being largely a rural state, is one of the highly endemic states for schistosomiasis in the northwest zone.

Understanding the risk factors associated with UGS and FGS is critical to developing effective intervention strategies for disease control and elimination. This study investigated the prevalence, risk factors, and public health implications of urogenital and female genital schistosomiasis in endemic communities of Northwest Nigeria. The findings will provide evidence-based recommendations for community-based interventions, improved diagnostic approaches, and targeted health policies to combat this neglected but important public health disease.

## MATERIALS AND METHODS

### Study Design

This is a cross-sectional, prospective and descriptive community-based study where data on the prevalence and risk factors of urinary and female genital schistosomiasis was collected across endemic communities in Jigawa State. The study described the prevalence of urinary and female genital schistosomiasis among reproductive women in relation to demographic and other risk factors such as age, occupation, educational qualification, drinking water source and water contact activities.

### Study Area

Jigawa State is in northern Nigeria, within the Sudan-Sahelian ecological zone, and is highly agrarian with rivers, irrigation schemes, and floodplains, which significantly influence disease patterns, particularly schistosomiasis. Jigawa State shares borders with Kano, Bauchi, Yobe, and Katsina States, as well as the Republic of Niger to the north. The selected LGAs (shown in Figure 1) are strategically distributed across the state as follows: Auyo LGA is located in the northeastern part of Jigawa, close to the Hadejia River Basin, Mallam Madori LGA is adjacent to Auyo, forming part of the Hadejia Valley Irrigation System. Ringim LGA is in central Jigawa, closer to Kano State, serving as a commercial hub, Taura LGA lies in southern Jigawa, characterized by extensive farmlands and floodplains, Dutse LGA (the State capital) is in south-central Jigawa, featuring urban development and administrative centers. And Buji LGA is in the southwestern part of Jigawa, relatively drier with less irrigation compared to other LGAs.

**FIGURE 1.** Map of Jigawa State showing the study sites.

The climate and weather of the study area is typical of Sudan Sahelian type. The rainy Season (May–October) has an annual rainfall varies from 600 mm in the north to over 1,000 mm in the south, with LGAs near Hadejia River (Auyo & Mallam Madori) receiving more rainfall. The dry Season (November–April). Harmattan winds bring cool, dry air from the Sahara, and temperatures range from 15°C to 40°C. Variation in temperatures with the hottest months (March–May) with often recorded temperatures above 38°C, while cooler months (December–February) drop below 20°C at night. Seasonal flooding along riverine areas (Auyo, Mallam Madori, Taura, and parts of Dutse) creates stagnant water bodies, supporting snail populations that host *Schistosoma* parasites. High temperatures accelerate evaporation, reducing some water sources in Buji and Ringim, leading to lower schistosomiasis transmission in these areas.

Jigawa State is mostly flat, with alluvial floodplains along the Hadejia River Basin in Auyo, Mallam Madori, and Taura Local government areas (LGAs). The major water bodies are Hadejia River, Kafin Hausa River, and Tiga Dam (Kano/Jigawa border). Flood-prone areas are Auyo and Mallam Madori, making them high-risk zones for schistosomiasis transmission. The dryland areas include Buji and parts of Ringim, where schistosomiasis prevalence is lower due to fewer stagnant water sources. Snail breeding grounds (Biomphalaria spp. & Bulinus spp.) are abundant along irrigation canals and swamps in Auyo, Mallam Madori, and Taura. Water contact activities (fishing, rice farming, bathing) increase infection risk in riverine communities.

The selected LGAs have a combined population of approximately 1.5 million people, mainly Hausa and Fulani ethnic groups (Table 1). High-density areas like Dutse and Ringim have better healthcare access. Nomadic Fulani communities in Buji and Taura have limited access to healthcare.

**Table 1.** Estimated population of the study Local Government Areas

Agriculture dominates the economy, with rice, millet, sorghum, wheat, vegetables, and livestock farming being the primary sources of livelihood. Irrigation farming is common in Auyo, Mallam Madori, and Taura, leading to frequent water contact and higher UGS/FGS risks. Ringim and Dutse are major commercial centers, with trade in livestock, textiles, and grains. Buji relies on rain-fed farming, with less irrigation, leading.

The distribution of health facilities shown below (Table 2) in the study sites reveals disparity among the six LGAs. Dutse has the best medical infrastructure, while Buji and Taura have the least access to healthcare. FGS cases are often misdiagnosed as STIs, requiring more trained gynecologists in local hospitals.

**Table 2.** Distribution of Health Facilities in the Study Sites

Although there is effort to curtail open defaecation in the state, the practice is common in rural areas. of the six LGAs, Ringim and Dutse have better WASH facilities, while some communities in Auyo and Mallam Madori rely on open water sources for drinking and domestic use. Limited boreholes and pipe-borne water in Buji and Taura increase exposure to contaminated water sources.

The six LGAs in Jigawa State display significant variations in geography, climate, economic activities, and healthcare access, all of which influence schistosomiasis transmission patterns. The Hadejia River Basin LGAs (Auyo, Mallam Madori, and Taura) remain high-risk zones, while Buji and Ringim have lower transmission rates due to fewer water contact opportunities.

### Selection of Study Sites

As shown in Figure 1, the study sites at each of the three senatorial districts were purposely selected based on the presence of irrigation projects and history of reported cases of Schistosomiasis by the State Ministry of health. In each Senatorial district two local government areas (LGAs) were selected. In Central Senatorial district the LGAs selected were Dutse and Buji, Northeast Mallam-Madori and Auyo and Northwest Ringim and Taura. In each of the selected LGAs, two communities were chosen, except in Dutse where three were purposely chosen based as shown in Table 3 below. In Auyo LGA, the communities selected were Gatafa and Kalgwai; in Malam-Madori LGA, Arki and Malam-Madori town; Ringim LGA, Majiya and Nahuce; Taura LGA, Yandutse and Shafar; Buji LGA, Ahoto and Gantsa; Dutse LGA, in Wurma, Warwade and Baranda.

**Table 3.** Selected communities from Each Senatorial Zone

### Sample Size Determination

The sample size calculation was based on the prevalence of a previous study on *Schistosoma haematobium* infection. Allowing for a 5% margin of sampling error and 95% confidence limit, sample size needed was determined by the formula described previously (34, 35).

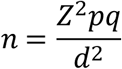

Where n = Sample size

Z = normal standard deviations, Z = 1.96 (statistic for a level of 95% confidence level)

P = expected prevalence or proportion (e.g. if 80 %; P= 0.80)

q = 1-P (1-0.80 = 0.79)

d = Degree of precision (e.g. if 5%, d = 0.05)

Thus, the minimal number of samples collected in the study where at least 80% power at 5% significance is 306 (including 20% drop out rate), but we collected 648 samples. The researcher doubles the estimate to strengthen the study and account for participant withdrawal or dropout. However, the sample size calculated was adjusted to 648 as the baseline sample size of the study to avoid bias in the selection of the participants from each of the two chosen villages in each local government. Auyo (166), Malam Madori (119), Dutse (68), Buji (119), Ringim (92), and Taura (84).

### Study Population

The study population comprises of 648 consented and assented participants’ female adults and girls’ residents between the ages of 5-49 years old. With clinical investigation of FGS there were further additional selections criteria including: inclusion and exclusion. Only those consented or assented studied participants that signed and agreed to participate in the study, among which included adult women, married and divorcée with age range (15-50 years) were recruited for both urinalysis and pelvic examination (colposcopy) of Female genital schistosomiasis (FGS) that had sexual activity (for easy speculum-aided gynaecological investigation for vaginal and cervix examination for any abnormalities. while girls between the age range of (5-14 years) were only recruited for urinalysis for presence of *Schistosoma.* And those living in the area for more than six months.

Those studied participants that did not signed, agreed and consented to participate in the study, pregnant women, women within two months of delivery, females on menstrual period / one week after menstrual period, women that have undergone hysterectomy and virgins were excluded (having declared no prior sexual activity were excluded since this does not allow the use of a speculum in the investigation. And those not living in the area for more than six months. Samples were collected from consented and assented participants in the studied area (two villages in each district with exception of Dutse that constituted three villages,

## METHOD OF DATA COLLECTION

### Community Sensitisation and Mobilization

Initially, community mobilization and sensitization were carried out with the assistance of Jigawa State Neglected Tropical Diseases/Eye Care Coordinator with corporation of village/ward heads, Ward Development Committee (WDC) and Voluntary community mobilizers in each community. Both the adult participants and the child’s guardian gave their written consent and assent after being informed about schistosomiasis and the purpose of the study’s objectives. Participants were informed that taking part in the study was voluntary and that they may discontinue at any time. Participants received full disclosure of the study’s objectives, their freedom to agree or decline, data privacy issues, and confidentiality concerns. Data was collected between March 15^Th^ 2021 and August 29^Th^ 2022.

### Sample Collection and Processing

After mobilization, urine samples were collected from consented and assented participants. Labelled, clean, wide mouth, screw cap, transparent containers were given to the participants for mid-day urine samples collected between 10am to 2pm. The urine sample collected were instantly observed for macrohaematuria and recorded the positive respondent, tested for microhaematuria and proteinuria were then transported in an ice container to the Biology Laboratory, Department of Biological Sciences and Federal University Dutse, Jigawa State, Nigeria. For microhaematuria 10ml urine samples was centrifuged at 5000 rpm for 5 minutes using sedimentation technique as described in previous studies (36) in which the supernatant was discarded using syringe and needle, and the sediment was transferred to a plain slide with cover slip and the sediment was examined initially at ×10 and then × 40 magnifications under the light microscope, to identify for the presence of *Schistosoma haematobium* ova which is characterized with a terminal spine. The eggs were counted and recorded as eggs/10 ml of urine. (37, 38) . Proteinuria was screened using 10 ml of urine sample with chemical reagent strips (Medi-Test Combi-10 manufactured by Machery-Hagel Duren, Germany)/ (Siemens Multistix 10 SG). The reagent end of the test strip was dipped into fresh, well-mixed uncentrifuged urine specimen for 40 seconds. After removal, then matched with the colour chart on the test strip container for detecting the presence of blood in urine value as described by the manufacturer. Anthropometric measurements such as height and weight were taken as described by (39). The body mass was measured in kilograms (kg) to the nearest 0.1kg using a digital floor weighing scale with participants without shoes. While a measuring tape was used to measure the height in meters (m).

### Clinical and Colposcopy Examination

Prior to the gynecological examination (colposcopy), the procedure involved in clinical examination was explained in detail to each consenting participant. Participants underwent pelvic examination performed by an experienced gynaecologist. The physical investigation was done using simple visual inspection with acetic acid (VIA) using World Health Organisation (40) guidelines. The method started with inspection of the external genitals vagina and vulva followed by the cervix using disposable speculum swab with 5ml of 5% acetic acid diluted with 95ml of distilled water, abnormal areas react with the acid and turn white (40, 41). With the aid of a disposable and transparent vaginal speculum, the appearance of the vulva/ vagina and cervix was noted and by means of a hand-held colposcope an Olympus OCS 500 colposcope with a mounted Olympus E420 (10 Mpx) single lens reflex (SLR) camera or a Leisegang colposcope with a mounted Canon EOS 650D (18 Mpx) SLR, the colposcopy examination was conducted. Any observed abnormalities or lesions were documented on paper by the investigator and later entered into electronic investigation forms by one data entry clerk, and if the cervix was visible through the colposcope, pictures of the images were captured before and after application of acetic acid via the computer screen.

Identification of the digital images for any abnormalities among reproductive age women in the studied area, cervico-vaginal surfaces were inspected according to WHO guidelines FGS pocket atlas, specifically notified clinical indication for FGS as indicated by the five clinically visible manifestations such as contact bleeding, yellow sandy patches, grainy sandy patches, rubbery papules and abnormal blood vessels (42).

Images captured were compared to the WHO FGS pocket atlas (42) to note significant consequences; these were subsequently saved in a coded database for internal validation through the blinded inspection of cervical pictures from photo colposcopy by some experienced health workers, using the FGS pocket atlas.

### Questionnaire Administration

Female research assistants were recruited and trained on administering a standardized questionnaire in a sensitive and culturally appropriate manner. They familiarized themselves with the questionnaire content and conducted interviews at the village health clinics. Informed consent or assent (age-appropriate) was obtained from each participant before proceeding. The prepared and validated questionnaire was used in collecting the following information:

i. Sociodemographic Age, residence, marital status, educational level, and occupation.
ii. Socioeconomic indicators: Household water source (borehole, stream, tap, well, piped water), frequency of contact with water open source, and the reason of contact or form activity (domestic chores, bathing).

### Ethical Clearance

Ethical clearance was obtained from the Federal University Dutse Research Ethics Committee (FUDREC) [Ref. Nos. FUDREC/2021/0011; and FUDREC/2021/1003], Federal University Dutse, Jigawa State. The research was also approved by the Jigawa State Health Research Ethics Committee, Jigawa State Ministry of Health (Ref. No. JHREC/2021/004; and JGHREC/2021/004). Each community’s district head was informed of the study’s purpose and provided with a copy of the state authorities’ clearance approval. More importantly, each participant signed a consent form.

## RESULTS AND DISCUSSION

### Results

A total of 648 females from six local government areas (LGAs) were enrolled into the study, details of the participants’ characteristics is presented in Table 4 (a & b). In the six LGAs, the population were; in Auyo (166; 25.6%), Buji (119; 18.4%), Malam Madori (119, 18.4%), Ringim (92; 14.2%), Taura (84; 12.9%) and Dutse (68; 10.5%). The participants’ ages ranged from 5 to 50 years, with a mean of 25.8 ± 10.2 years and a median of 25 years (interquartile range: 19–32 years). The predominant age group was 20 – 29 years old (36.7%) followed by 30 – 39 years old (25.5%). The least represented group were girls between the ages of 5 – 9 years old (3.9%). The majority of the overall study participants were petty traders (62.6%), mainly engage in small businesses like selling soya beans balls (Awara), beans cake etc. Some participants solely depend on their spouses (housewives, 18.7%), fewer were students (12.6%), and Some Were Hawkers (4.9%). Across all the LGAs majority were petty traders, but in Auyo and Dutse, there were more students (15.7% and 11.8%) than House wives (13.9% and 10.3%) respectively. Overall, the participant’s spouse/parent were mostly famers (50.5%) and petty traders (44.9%), few were civil servants (4.6%). The trend was same in all LGAs except in Taura and Dutse whom were more petty traders than farmers (42.9% vs 53.6%; 45.6% vs 52.9%) respectively.

**Table 4.** General Characteristics of the study participants

About two-thirds (65.6%) of the participants had no formal education, followed by those with primary (19.4%) and secondary (13.6%) as highest level attained. By the LGAs, in M. madori, more participants attained secondary level of education (25.2%) than those with primary (13.4%). Most of those with tertiary education were from M. madori (5 out of 9 total). Similarly, the spouse/parent’s educational status showed that majority had no formal education (71.6%) but among those with formal education more attained secondary level (13.5%) followed by those with primary (8.5%) and least were those with tertiary level of education (6.4%). Same trend was seen in all the LGAs.

### Prevalence of Urogenital Schistosomiasis

Of the 648 participants screened, 88 were positive for urogenital schistosomiasis (UGS) infection, resulting in an overall prevalence of 13.6% (95% CI: 11.0, 16.5%). The prevalence of UGS infection differed significantly across the study locations: χ2 = 31.3, *p* < 0.001. As shown in Figure 2, Auyo had the highest prevalence of UGS (6.3%). A moderate prevalence was seen at Mallam-Madori (2.2%) and Taura (1.9%) and was significantly higher than the prevalence at Dutse (1.5%) and Ringim (1.1%). The least prevalence was recorded at Buji (0.6%). The distribution of UGS prevalence across the villages sampled, depending on the village, ranged from 0% in Kalgwai and Gantsa to 35% in Gatafa. The prevalence (and its 95% CI) of UGS in all the sampled villages in each location is shown in Table 5. Depending on the village, the prevalence ranged from 35% in Gatafa to no infection (0%) in Kalgwai and Gantsa.

**FIGURE 2.** Prevalence of urinogenital and female genital schistosomiasis in the six local government areas

**Table 5:** Prevalence of Urogenital and Female Genital Schistosomiasis by Study Locations Prevalence of Female Genital Schistosomiasis.

Among the eighty-four (84) participants with Schistosome eggs detected in the urine sample, 21 (25%) had heavy infection (≥ 50 eggs/ml of urine) while 63 (75%) had light infection (< 50 eggs/ml of urine). Four samples, though without eggs, were positive for both hematuria and microhaematuria. The Chi-square test (χ2 = 13.31, *p* = 0.02) indicated that the proportion of participants with heavy infection (≥ 50 eggs/ml of urine) was significantly higher in Ringim (4/7; 57.1%) and Taura (6/12; 50%) compared to Dutse (3/10; 30%) and Auyo (8/41; 19.5%). All cases of UGS infection in Buji (n = 4) and Malam Madori (n = 10) were classified as light infections, with 2 – 8 eggs/ml of urine.

Of the 648 participants, 606 (93.5%) consented to gynecological examination as shown in Figure 2, the overall prevalence of female genital schistosomiasis (FGS) in the study sample was 152 (25.1%). This means that one in five females was positive for at least one of the six clinically visible manifestations: contact bleeding, pre-contact bleeding, yellow sandy patches, grainy sandy patches, rubbery papules and abnormal blood vessels. In the study population, the prevalence lies somewhere between 21.7% and 28.7% (95% confidence interval). The prevalence (and its 95% CI) of FGS in all sampled villages in each location is shown in Table 5. Depending on the village, the prevalence ranged from 8.5% in Kalgwai to 61.5% in Wurma. In addition, the two villages at Buji recorded high FGS prevalence, almost twice the overall prevalence: Ahoto (43.9%) and Gantsa (45.2%).

The difference in FGS prevalence across the study locations was statistically significant: χ2 = 38.125, *p* < 0.001. The prevalence at Mallam-Madori (21.8%), Taura (21.4%) and Ringim (20.6%) was comparatively similar and significantly lower compared to Dutse (38.7%) and Buji (44.5%). The location with the lowest prevalence of FGS was Auyo (14.9%).

### Factors Associated with Urogenital and Female Genital Schistosomiasis

Factors such as water contact activities, toilet at home and urinating outside were examined for significant association with urogenital and female genital schistosomiasis infection both at the bivariate and multivariate level (adjusting for location and age). Table 6 shows the crude and adjusted odds ratio for factors associated with urogenital schistosomiasis (UGS). At the bivariate level, water contact activities and urinating outside were significantly associated with UGS infection. These significant factors from the bivariate analysis were inserted into a multivariate model to adjust for the effect of age and location. At the multivariate level, both factors failed to attain statistical significance. The result suggested that participants engaged in swimming were 1.7 (*p* = 0.30) times more likely to have UGS infection compared to those with no contact with waterbodies. The empirical data shows that 30.6% of participants engaged in swimming were infected with UGS compared to 12% of those without contact with water bodies or whose water contact activities were domestic in nature (Table 6). Regarding location, compared to Buji LGA with the lowest infection rate, the likelihood of UGS infection was 9.3 (*p* < 0.001) times as high in Auyo, 4.5 (*p* = 0.02) times as high in Dutse, 4.7 (*p* = 0.01) at Taura, 4.2 (*p* = 0.01) times higher in Malam Madori and 2.4 (*p* = 0.16) times as high in Ringim. Additionally, the odds of UGS infection decreased with age, with the highest likelihood seen in 10 – 14-year-olds (odds ratio = 5.56, *p* = 0.006) and 15 – 19 years olds (odds ratio = 3.34, *p* = 0.05).

**Table 6.** Bivariate and Multivariate Analysis of Factors Associated with Urogenital Schistosomiasis

Concerning female genital schistosomiasis (FGS) (Table 7), at the multivariate level, water contact activities and urinating outside were not statistically significant predictors of FGS infection. Rather, location and age were independently and significantly associated with FGS infection. Auyo LGA had the lowest infection rate (14.9%). Compared to Auyo, the likelihood (odds ratio) of FGS infection was 3.9 times as high in Buji (44.5%; *p* < 0.001) and 2.3 times as high in Dutse (38.7%, *p* = 0.05). For the other study locations, the odds ratio did not attain statistical significance. The result also suggested the likelihood of FGS infection increased with age, that is, older women were more likely to have FGS compared to younger girls (10 – 14 years) (see Table 7).

**Table 7.** Bivariate and Multivariate Analysis of Factors Associated with Female Genital Schistosomiasis

## Discussion

An overall urogenital schistosomiasis (UGS) prevalence of 13.6% among 648 female participants communities in Jigawa State, Nigeria, posed a significant public health concern. The variation in prevalence across the LGAs (χ² = 31.3, p < 0.001) could be due to localized transmission dynamics, likely influenced by differences in water contact behavior, sanitation, and control interventions (43). The highest UGS prevalence recorded in Auyo (6.3%), moderate in Mallam-Madori (2.2%) and Taura (1.9%), while Buji had the lowest (0.6%), This geographical differences in prevalence could be explained in the context of previous findings with previous studies, which shows that schistosomiasis transmission is often clustered around water sources where intermediate host snails thrive (23, 27). Auyo being an LGA with highest prevalence correlates with the area being a rice growing area with its vast irrigation scheme.

The variation in intensity of infection within the study population where a quarter (25%) of the infected participants had heavy infections (≥ 50 eggs/ml of urine), with significantly higher rates in Ringim (57.1%) and Taura (50%) compared to Auyo (19.5%) and Dutse (30%) (χ² = 13.31, p = 0.02) revealed that certain LGAs experienced higher rates of active transmission or reinfection cycles. This, according to (20, 24), could be due to poor sanitation, frequent water contact, and limited access to treatment. The presence of haematuria and microhematuria in four participants without detectable eggs suggests potentially early-stage infections or false-negative microscopy results, highlighting the need for more sensitive diagnostic tools such as PCR-based techniques (15).

All UGS cases in Buji and Mallam-Madori were light infections (2–8 eggs/ml) suggesting low transmission intensity in these areas. This could be accounted in terms of differences in host immunity and water exposure patterns (1). Moreover, in these communities, there is less irrigation or all-year-round farming activities when compared to other study sites such as Auyo, Ringim & Taura. The study’s findings reinforce the importance of targeted intervention strategies, including mass drug administration (MDA), improved water sanitation, and behavioral change campaigns, particularly in high-burden areas like Auyo, Ringim, and Taura (17, 25).

The high prevalence (25.1%) of Female Genital Schistosomiasis (FGS) among participants who underwent gynecological examination shows that a significant number of infected UGS women go on to manifest FGS. This could be due to lack of prompt treatment or its failure and continuous reinfection. This indicates that FGS poses a significant public health burden in the study area. The wide variation in FGS prevalence across locations (χ² = 38.125, p < 0.001) suggests localized transmission dynamics due to various risk factors, including water contact behavior, access to healthcare, and diagnostic awareness previously reported by (44, 23). The highest prevalence recorded in Wurma (61.5%), and villages of Buji LGA (Ahoto: 43.9%, and Gantsa: 45.2%) also experiencing disproportionately high FGS rates. These findings agree with previous studies, which highlight that FGS is particularly prevalent in rural communities with poor sanitation and frequent exposure to contaminated water sources (24, 20).

The significant difference in FGS prevalence across LGAs further underscores the heterogeneous nature of schistosomiasis transmission. The lower prevalence in Auyo (14.9%) and Mallam-Madori (21.8%) compared to Dutse (38.7%) and Buji (44.5%) may be attributed to differences in exposure to infected water bodies, health-seeking behavior, and the effectiveness of prior control measures which is agreement with the works of (7), & (15). In fact, the lower prevalence in Auyo could be largely due to the effectiveness of control measures, especially treatment with praziquantel at early stages despite the LGA accounting for the highest UGS infection.

The strong association between FGS and gynecological symptoms, such as contact bleeding, sandy patches, and abnormal blood vessels, highlights the urgent need for improved diagnostic and treatment strategies. Studies suggest that FGS is often misdiagnosed as other reproductive tract infections, leading to inadequate treatment and increased risk of complications, including infertility and susceptibility to HIV and HPV (10, 4).

The potential risk factors for urogenital schistosomiasis (UGS) and female genital schistosomiasis (FGS), identified age and location as the most significant predictors of infection. However, water contact activities and urinating outside were associated with UGS at the bivariate level, they lost significance in the multivariate analysis, suggesting that age and geographic location played a stronger role in infection risk which is in agreement with position of (4, 23).

UGS infection was highest in Auyo (OR = 9.3, p < 0.001), followed by Dutse (OR = 4.5, p = 0.02), Taura (OR = 4.7, p = 0.01), and Mallam Madori (OR = 4.2, p = 0.01), with Buji LGA having the lowest risk. This geographic disparity suggests localised transmission patterns, likely influenced by differing levels of exposure to infected water sources, sanitation infrastructure, and past control interventions (20, 17).

Age was a strong independent predictor of UGS infection, with adolescents (10–14 years) having the highest risk (OR = 5.56, p = 0.006), followed by 15–19-year-olds (OR = 3.34, p = 0.05). This agrees with existing literature indicating that school-aged children and adolescents have the highest infection rates due to frequent contact with contaminated water during recreational and domestic activities (1, 24).

Similarly, FGS prevalence was strongly associated with location and age. Compared to Auyo (14.9% prevalence), women in Buji had a significantly higher likelihood of FGS infection (OR = 3.9, p < 0.001), followed by Dutse (OR = 2.3, p = 0.05). The association between FGS and age suggests that older women are at greater risk, possibly due to longer exposure to infected water sources and cumulative parasite burden over time (15, 10).

## Conclusion and Recommendations

### Conclusion

An overall UGS prevalence of 13.6% was recorded in this study, while that of FGS was 25.1%. The high variability in prevalence across different study locations is attributed to environmental, behavioral, and socio-economic factors that influence transmission dynamics. Auyo LGA had the highest prevalence of UGS (6.3%), while Buji recorded the lowest (0.6%). On the other hand, Buji and Dutse had the highest FGS prevalence (44.5% and 38.7%, respectively), with Auyo having the lowest (14.9%).

Age and location were found to be independent predictors of UGS and FGS infections. However, water contact activities and urinating outside did not remain significant after adjusting for these factors. The highest risk group for UGS was adolescents (10–14 years old), while older women had the highest risk for FGS, suggesting cumulative exposure over time. Severe cases (≥50 eggs/ml urine) encountered in certain LGAs, most especially Ringim (57.1%) and Taura (50%), could be potential hotspots for active transmission.

### Recommendations

1. Strengthening and sustaining Mass Drug Administration (MDA) campaigns to cover high-risk populations, particularly adolescents and older women in endemic areas. In the context of findings from this study, there should be consistent and widespread distribution of praziquantel, especially in high-prevalence LGAs like Auyo, Buji, and Dutse.
2. There should be efforts on the part government to increase access to clean and safe water sources to reduce dependency on contaminated water bodies. The construction and use of improved latrines to further minimize open defaecation (ODF) that could drastically reduce environmental contamination there interrupting transmission. This should be complemented by implementing hygiene education programs to raise awareness about the risks of water contact activities such as swimming and washing in infested water sources.
3. Community-Based Health Education and Awareness Campaigns should be enhanced through targeted awareness programs in schools, markets, and religious centers on the symptoms, transmission, and prevention of UGS and FGS. In addition, the engagement of traditional and religious leaders in behavioral change campaigns to promote early health-seeking behavior. There should also be radio, television, and social media campaigns to disseminate information on FGS and its association with infertility, HIV risk, and reproductive health complications.
4. Strengthening diagnosis and treatment services should be pursued by training healthcare workers, particularly gynecologists and primary healthcare providers, to recognize and diagnose FGS since it is often misdiagnosed as other reproductive health conditions. There is a need to expand diagnostic capacity, including the use of microscopy, point-of-care rapid tests, and molecular diagnostics, especially in rural health facilities.
5. Conducting further research and surveillance by establishing longitudinal studies to monitor schistosomiasis prevalence trends and assess the impact of ongoing interventions. There is also a strong need to study local transmission dynamics, including snail vector distribution and human-water contact behaviors, to refine control strategies. This will guide the development of an integrated disease mapping programs to guide resource allocation for schistosomiasis control efforts.

## Data Availability

All data regarding the manuscript has been included in the manuscript submission here.

